# The ACROSSS-study – the role of sleep characteristics before and after a cardiovascular event. Rationale and study design

**DOI:** 10.64898/2026.11.18.26368283

**Authors:** Anna-Therese Klötzsch, Tobias Kühl, Martin Christ, Birgit Hailer, Benedikt Frank, Martin Köhrmann, Christoph Kleinschnitz, Oliver Bruder, Andreas Stang

## Abstract

Studies of myocardial infarction and stroke showed a diurnal variation with peaks in the morning and afternoon. So far, night sleep or napping have not been investigated as triggers for cardiovascular events via a case-crossover study. Therefore, the main question of the ACROSSS (**<u>A</u>**cute **<u>C</u>**o**<u>RO</u>**nary **<u>S</u>**yndrome, **<u>S</u>**troke and **<u>S</u>**leep) study is how much the occurrence of a myocardial infarction/stroke/transient ischemic attack is associated with taking a nap shortly before (topic 1, case-crossover study, 656 patients needed). Moreover, the study aims at assessing the incidence of sleep disturbances three and twelve months after a myocardial infarction/stroke/transient ischemic attack (topic 2, follow-up study, 400 patients needed). Recruitment of patients is carried out in four German hospitals in Essen and Bottrop, North Rhine-Westphalia. The examination program includes the case-crossover questionnaire on potential acute triggers of cardiovascular events, a sleep questionnaire with validated instruments (Pittsburgh Sleep Quality Index, Epworth Sleepiness Scale, etc.) and a questionnaire on health status and lifestyle habits. Moreover, eligible patients are screened for sleep apnea and during the follow-up sleep is measured by actigraphy. The combination of sleep-related and clinical data offers a deeper understanding of how sleep affects the acute onset of a cardiovascular event and recovery afterwards.

## Introduction

According to the World Health Organization, ischemic heart disease and stroke were still the most frequent causes of death worldwide in 2019 [1]. In the Global Burden of Disease Study 2019, the prevalence of ischemic heart disease was 2421 per 100,000 persons and the incidence rate was 262.4 per 100,000 person-years [2]. The prevalence for stroke was 1240 per 100,000 persons and the incidence rate was 150.8 per 100,000 person-years. Research on cardiovascular diseases has been ongoing for many decades including the analysis of risk factors [3]. The effects of diabetes, hypertension, dyslipidemia and smoking have been well studied and are taken into account in public health strategies to prevent cardiovascular diseases [4]. These risk factors have a long-term effect on the cardiovascular system and over time cause endothelial cell dysfunction and atherosclerosis [5]. However, acute triggers of cardiovascular events (myocardial infarction, stroke and transient ischemic attack [TIA]) usually have a short latency, this means a short time between occurrence of the trigger and occurrence of the effect, such as heavy physical exertion, stress or anger, certain drug uses and respiratory infections [6]. In the search for other possible triggers, it has been noted that acute myocardial infarctions occur more frequently in the morning and afternoon [7, 8]. It is therefore assumed that not only night sleep but also daytime sleep (napping) may be a relevant acute trigger for myocardial infarction and stroke, since many people at an advanced age also take a nap [9]. So far, only the association between night sleep and the occurrence of myocardial infarction a few hours after waking up has been investigated. To our knowledge, no studies have yet been conducted on napping as a potential trigger for myocardial infarction. Due to the similarities of the relevant risk factors of myocardial infarction and stroke, the study is conducted in both myocardial infarction patients and stroke patients. Therefore, the main study question is how much the occurrence of a myocardial infarction/stroke/TIA is associated with napping shortly before the event.

In addition to identifying possible triggers for myocardial infarction and stroke/TIA, it is also important to continue monitoring patients after a cardiovascular event and to support patient recovery. A first-time myocardial infarction or stroke is a life-changing event for most people. Many patients suffer from depression, anxiety, exhaustion or disturbed sleep afterwards [10,11]. In addition to the negative effects of sleep disturbances on quality of life, Alcántara et al. found a 50% higher rate of a new myocardial infarction within the next year in patients who slept less than 7 hours (hazard ratio=1.52; 95% CI: 1.06-2.18) [12]. Previous studies have found a prevalence of 21 to 33% for sleep disturbances after a myocardial infarction [10,13–15]. In a systematic review, a disturbed sleep architecture measured by polysomnography or actigraphy was found after acute coronary syndrome [16]. However, these study results were only based on small case numbers or lacked data on sleep disturbances before the myocardial infarction. Studies on the incidence of sleep disturbances or naps after a first myocardial infarction or stroke are still pending. Therefore, the second study aim is to assess the incidence of sleep disturbances three and twelve months after a myocardial infarction or stroke/TIA.

### Main research questions

#### Topic 1

In a case-crossover study, we want to estimate the strength of association between the time of occurrence of myocardial infarction/stroke/TIA and a nap taken shortly before. We also want to investigate other triggers for the occurrence of myocardial infarction/stroke/TIA such as characteristics of night sleep and the intake of non-steroidal anti-inflammatory drugs (NSAIDs) during respiratory tract infections.

#### Topic 2

We want to determine the incidence of sleep behavior changes, e.g. taking a nap, changing the nap duration as well as sleep disturbances in patients with first-time myocardial infarction/stroke or after any TIA 3 and 12 months after the index event.

### Study design

To answer the main research questions, two different study types are combined in a joint study. Topic 1 is addressed by a case-crossover study (**figure 1**). This is a relatively new methodological approach, which has greatly advanced the research of disease triggers. A case-crossover study is particularly suitable if short-term effects of temporary exposures (triggers) are to be investigated. The disease of interest must have a clear onset [17, 18]. Due to their sudden onset, myocardial infarctions and strokes/TIAs can be investigated well by means of case-crossover studies. If the exposure has a short-term effect, the duration of the exposure is limited to certain hours of the day and ends with the onset of the disease (case time interval). Among the same patients, this time interval is used to compare the hours before the event with a control period, where no event occurred (control time interval). The cases serve as their own controls and no additional control group is necessary. In the ACROSSS study, patients are questioned retrospectively during their hospital stay. Patients are asked if they experienced the triggers on the day of the event and one day before the event as well as the usual frequency of the trigger.

**figure 1.**
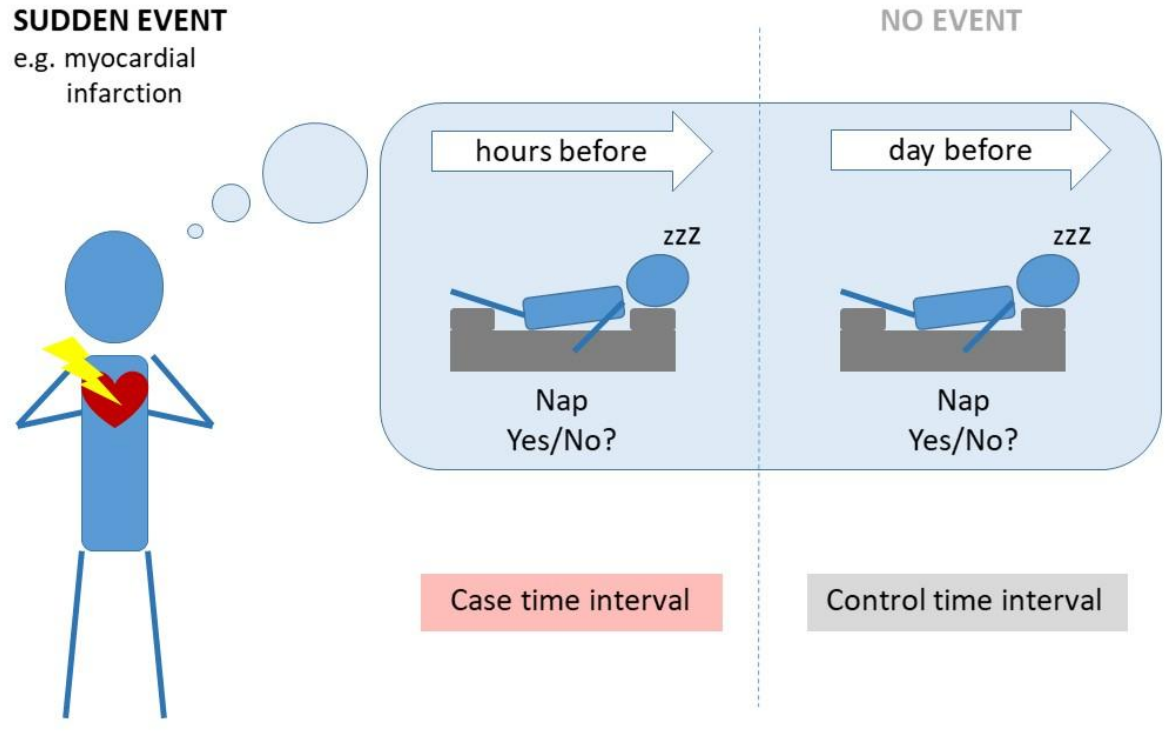
principles of a case-crossover study using the example of napping as potential trigger of myocardial infarction/stroke/TIA

Topic 2 is addressed by an incidence study (longitudinal study, follow-up study). The observation period is 12 months with an initial follow-up study 3 months after the myocardial infarction/stroke/TIA. The identical patients of the case-crossover study serve as patients for the incidence study.

## Methods

### Study setting and eligibility criteria

All patients with acute myocardial infarction or stroke/TIA who are admitted to the cardiologic wards of the Elisabeth Hospital, the Catholic Hospital Essen and the Knappschaftskrankenhaus Bottrop or the neurological ward of the University Hospital Essen and who meet the inclusion criteria are eligible for study participation.

Patients are eligible if they are at least 18 years old and if they are hospitalized for the treatment of a first-time acute myocardial infarction (ICD-10: I21) or first-time stroke or any TIA (ICD-10: I63, G45.8, G45.9). Patients with myocardial infarction must not present a leading heart valve vitium. Furthermore, patients are not eligible if they are not able to give informed consent or if they suffer from aphasia or cognitive impairment because of dementia, mentally confusion or disorientation.

### Timeline and examination program

The study personnel checks the newly admitted patients regarding the inclusion and exclusion criteria (**figure 2**). Eligible patients are informed about the study and have to give written informed consent to participate in the study. Afterwards the study personnel hands in the questionnaires to the patient and explains the content and structure (**table 1**). If the participants are not physically able to fill out the questionnaires, the study personnel conducts the questionnaires as standardized interview. In addition, for a fraction of patients, the study program includes a screening for sleep apnea via the device ApneaLink™Air or SOMNOtouch™RESP eco for one night in the hospital. The study personnel collects the questionnaires and the device on the following days. In certain circumstances, participants finish the questionnaires at home and send them back via mail.

**Table 1.** Examination program of the ACROSSS study, Germany, from 2018 until 2023.

| **Study element** | **Time point** | **Used in topic** |
|---|---|---|
| Case-crossover questionnaire | T1 | 1 |
| Questionnaire on sleep characteristics and sleep habits during the last 4 weeks | T1, T2, T3 | 1, 2 |
| Confounder questionnaire | T1 | 1, 2 |
| Sleep apnea screening (ApneaLink™Air or SOMNOtouch™RESP eco) | T1 | 2 |
| Objective sleep measurement (SOMNOwatch™plus) | T2, T3 | 2 |
| Medication (prescribed and over-the-counter) | T2, T3 | 2 |
| Follow-up questionnaire | T2, T3 | 2 |

**Figure 2.**
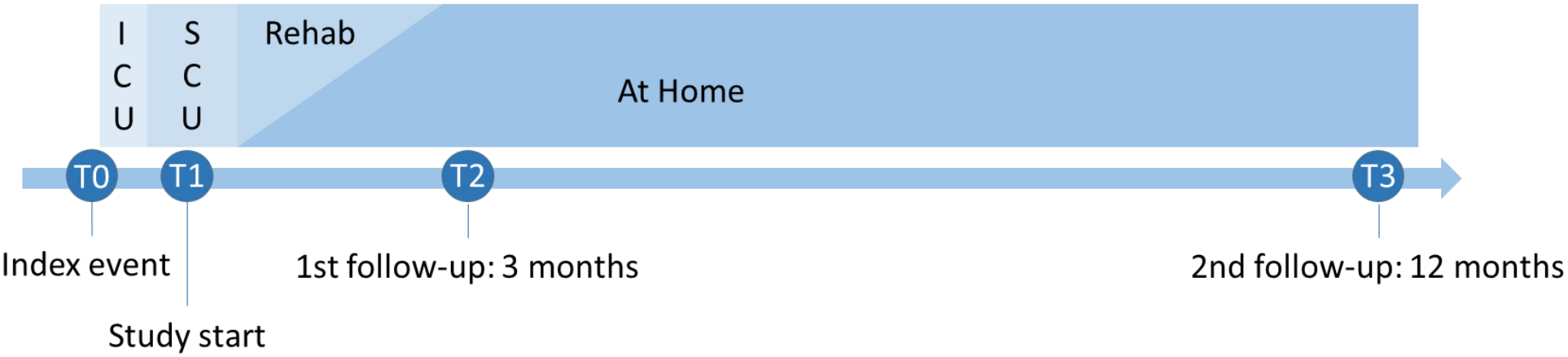
Study schedule of the ACROSSS study, Germany, from 2018 until 2023. ICU…intensive care unit, SCU…specialized care unit, Rehab…Rehabilitation

Afterwards the sleep apnea measurement is controlled for quality and plausibility and is analyzed at the sleep laboratory of the Ruhrlandklinik Essen. If the measurement is of good quality and the Epworth Sleepiness Scale is filled out completely, the study personnel sends the results of the sleep apnea screening to the participants.

After ten weeks, the study personnel starts contacting the participants for the first follow-up examination as part of the incidence study. Only patients living in Essen and the surrounding cities or communities are considered for the home visits within the incidence study. As an alternative to the home visit, patients are offered to come to the study center or to participate with a reduced examination program via mail. The complete examination program includes an activity measurement with the device SOMNOwatch™plus for 24 hours with the additional option of a single-lead electrocardiogram and flow detection during nighttime as well as questionnaires on sleep behavior, medication, and vital status. The reduced examination program includes the questionnaires as well as the activity measurement via SOMNOwatch™plus but without electrocardiogram or flow detection. The questionnaires are send to all patients via mail regardless of their place of residence.

The second follow-up examination twelve months after the myocardial infarction or stroke/TIA is organized in the same way. During times of high incidence for SARS-CoV-2, the patients are offered only the reduced examination program of the follow-up via mail. After five years of follow-up, there will be a request at the Residents’ Registration Office regarding the vital status of the patients.

### Questionnaires

#### Case-crossover questionnaire

The case-crossover questionnaire is available in two versions, one for the myocardial infarction patients and one for the stroke/TIA patients. At first, patients are asked about typical symptoms for these events and are instructed to indicate date and time of onset for each of them. Typical symptoms for stroke are sudden asymmetry of the face; pelt-like or numb feeling in arms or legs; loss of speech or difficulty understanding speech; impaired vision; unexplained dizziness, sudden tendency to fall; sudden disorientation [20]. Typical symptoms for myocardial infarction are severe pain in the upper body or in adjacent areas; massive feeling of tightness in the chest; severe burning; nausea, vomiting, shortness of breath, pain in the upper abdomen; anxiety sweat with cold pale skin [21]. After the questions on typical symptoms, both patient groups are asked the same questions on potential triggers, like lifestyle habits or emotions, regarding the hours before the event. Based on a literature research, we included the following potential triggers that seemed relevant for our patient cohort: stress, heavy meals, coffee consumption, alcohol consumption, use of analgesics, heavy physical activity, extraordinary events (e.g. championship final), respiratory infection, midday nap, sleeping times, special emotional situation (e.g. birth of a child, promotion or conflicts at work or in private life)[22–28]. If available, the same questionnaire is handed out to the partner or a close relative of the patient to verify or to complement the answers of the patients.

#### Sleep questionnaire

The sleep questionnaire consists of several validated instruments. To quantify the overall sleep quality of the patients, we use the Pittsburg Sleep Quality Index (PSQI) with a range from 0-21 points, with higher values indicating a lower sleep quality [29]. Moreover, the PSQI offers the analysis of the subcomponents subjective sleep quality, sleep latency, sleep duration, sleep efficiency, sleep disorders, use of sleeping pills and daytime sleepiness. The items on sleep-disordered breathing are included in the case-crossover questionnaire of the patient’s partner. To create separate items for difficulties maintaining sleep and early morning arousal, we split the original question in two items. Because many studies reported a poor sleep quality of patients during a hospital stay, we also include the German version of the Richards-Campbell Sleep Questionnaire [30]. In addition to the screening for sleep apnea, we use the Berlin questionnaire, which is a validated instrument to detect sleep apnea [31]. Because sleep apnea is associated with daytime sleepiness, we use the Epworth Sleepiness Scale as an instrument to rate daytime sleepiness as well as a necessary information to detect sleep apnea via the screening device ApneaLink™Air [32] or SOMNOtouch™RESP eco. The questions on napping were not taken from a validated questionnaire, but have been used in the Heinz Nixdorf Recall Study and the German National Cohort as well [33, 34]. The questions were “How often do you usually take a nap?” with the possible responses “never, less than once a week, 1 to 4 times a week, 5 to 6 times a week, every day” and “How many hours do you usually take a nap?”.

#### Confounder questionnaire

When planning the study, we did a literature search and used directed acyclic graphs to identify all confounders that are relevant for our main research questions [35]. The covariate-related questionnaire includes only confounders that were part of the minimally sufficient adjustment set or that are necessary for stratification. Besides information on social status, education level, marital status, employment situation and abilities of daily life, we ask about risk factors for cardiovascular diseases and poor sleep quality, namely smoking status, hypertension, BMI, dyslipidemia, noise level at residence, family history of cardiovascular disease, depressed mood and stress level. Relevant comorbidities are subjective health status, chronic obstructive pulmonary disease, diabetes mellitus, anemia and inflammatory joint disease. In addition, this information will be compared to laboratory measures and medical history from the hospital discharge letter.

#### Follow-up questionnaires

During follow-up after three and twelve months, the participants receive the same sleep questionnaire as during the baseline examination. The questions on depressive mood, abilities of daily life and subjective health are also repeatedly asked. Moreover, the participants answer questions about health events or hospitalizations in the meantime. In addition, the current medication is requested in a separate questionnaire.

### Devices

Sleep apnea is a risk factor for cardiovascular events and is highly correlated with sleep disturbances [36]. To separate the different effects, we have to assess if patients suffer from sleep apnea. For the detection of sleep apnea, we use a mobile screening device called ApneaLink™Air by Resmed or SOMNOtouch™RESP eco by SOMNOmedics GmbH. This device records nasal airflow via a nasal cannula, oxygen desaturation via finger sensor and the respiratory effort via a sensor at the chest strap. In a validation study, the device showed a high sensitivity and specificity for detecting an apnoe-hypopnoe-index ≥ 15 compared to polysomnography also in a home setting [37]. Moreover, the device was already used in the Heinz Nixdorf Recall Study, another study within the same region and with a similar age range [38].

Despite the several validated items included in the sleep questionnaire, we also use a device to get objective data on sleep. Because a polysomnography is too complex and expensive, we use the actigraph SOMNOwatch™ plus to assess sleep during the follow-up after 3 and 12 months. The SOMNOwatch™plus is worn on the wrist of the non-dominant hand and measures acceleration in three axes with a sampling rate of 128 Hz. The device is also combined with a 1-channel-electrocardiogramm with a sampling rate of 256 Hz. In order to get a good quality of the signal, the study personnel must place 3 adhesive electrodes on the patient’s upper body. During sleep, the patient is instructed to use a nasal cannula to measure the nasal airflow with a sampling rate of 256 Hz. The SOMNOwatch™plus showed good agreement with polysomnography regarding total sleep time, wake after sleep onset and sleep efficiency and is also used in the German National Cohort [39].

### Outcome measures

The case definition of the case-crossover study (Topic 1) refers to patients with acute myocardial infarction, stroke or TIA. Therefore, the study personnel checks the admission and discharge diagnoses for the corresponding ICD10-Codes: I21 for acute myocardial infarction, I63 for stroke and G45 for TIA (see confirmation of diagnosis and eligibility criteria). The primary outcome of the incidence study (Topic 2) was the incidence of sleep disturbances taken from the follow-up questionnaires after 3 and 12 months. The incidence of other sleep characteristics served as secondary outcome measures of Topic 2 (**Table 2**).

**Table 2.**
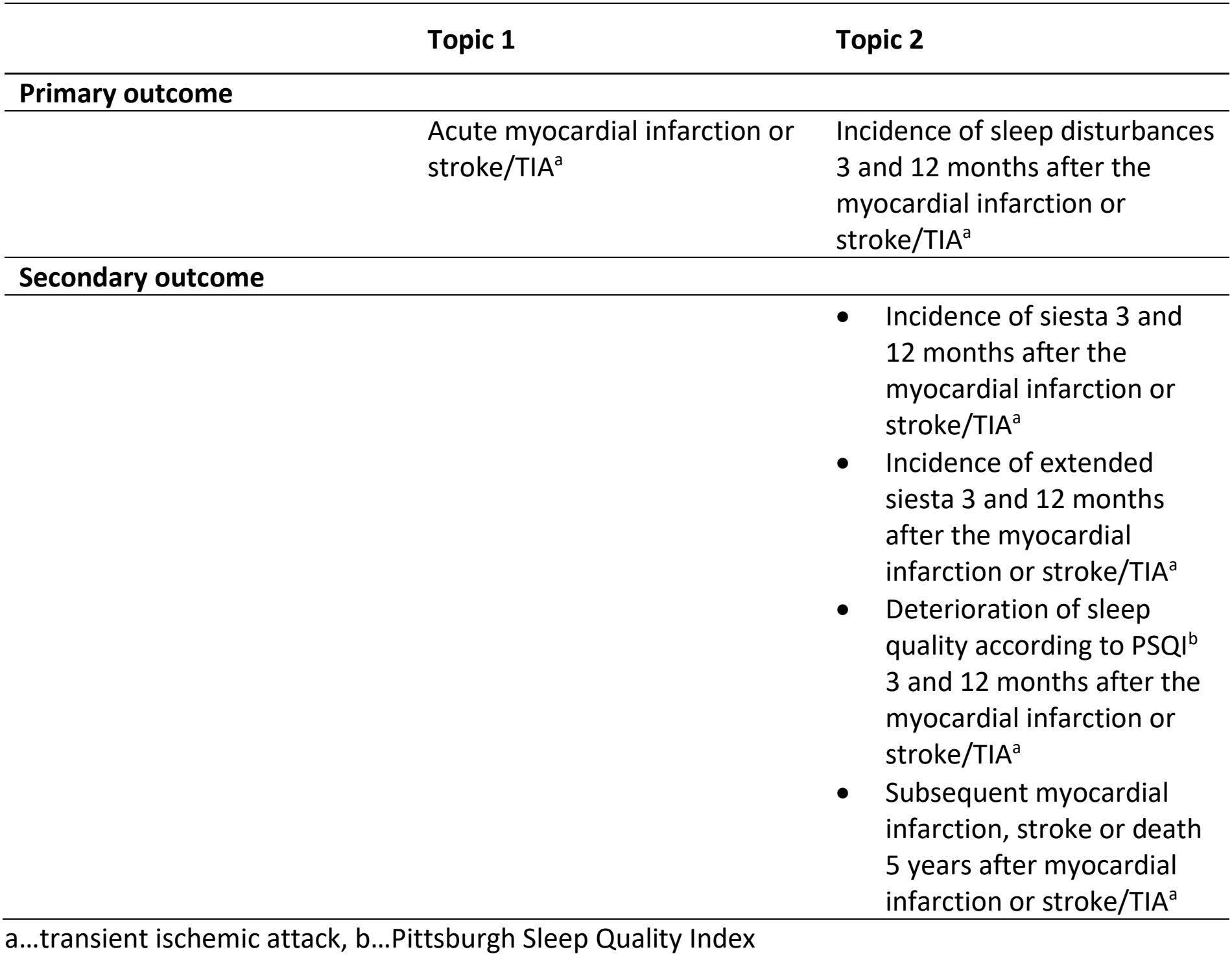
Primary and secondary outcome measures for both research topics of the ACROSSS-study.

|  | **Topic 1** | **Topic 2** |
|---|---|---|
| **Primary outcome** |  |  |
|  | Acute myocardial infarction or stroke/TIA<sup>a</sup> | Incidence of sleep disturbances 3 and 12 months after the myocardial infarction or stroke/TIA<sup>a</sup> |
| **Secondary outcome** |  |  |
|  |  | • Incidence of siesta 3 and 12 months after the myocardial infarction or stroke/TIA<sup>a</sup><br>• Incidence of extended siesta 3 and 12 months after the myocardial infarction or stroke/TIA<sup>a</sup><br>• Deterioration of sleep quality according to PSQI<sup>b</sup> 3 and 12 months after the myocardial infarction or stroke/TIA<sup>a</sup><br>• Subsequent myocardial infarction, stroke or death 5 years after myocardial infarction or stroke/TIA<sup>a</sup> |

a…transient ischemic attack, b…Pittsburgh Sleep Quality Index

### Sample size calculation

The following assumptions were made for the case-crossover study: two-sided test, alpha level of 0.05 and a power of 0.80. The difference between the proportions of the discordant pairs was assumed to be 0.10. The proportion of exposed persons during the control time window was set to 0.16 according to the napping prevalence in a local cohort study [19]. Under these assumptions, 328 patients are required for the case-crossover study. The sample size calculation for the incidence study considered the width of the 95% confidence interval. The cumulative incidence for sleep disturbances was assumed to range between 10% and 40%. With a probability of 99%, the half-width of the confidence interval would be 7%. Under these assumptions, 200 patients are required for the incidence study. As these sample sizes are necessary for myocardial infarction patients and stroke/TIA patients separately, a total sample size of 656 patients is needed for the case-crossover study and 400 patients for the incidence study. All power calculations were done with SAS (PROC POWER).

### Statistical analysis

After the definition of exposure and control time windows specifically for each exposure, McNemar odds ratios with 95% confidence intervals will be estimated. Because in a case-crossover study participants are both cases and controls, it is not necessary to control for time-invariant confounding. For the second research question, we will estimate prevalence and incidence for sleep characteristics including sleep disturbances, siesta and changes in sleep scores. All statistical analyses will be conducted with the latest available version of SAS®.

### Recruitment

Staff members screen newly admitted patients on five days a week in every study center. If informed consent was provided but questionnaires were not completed after six weeks, patients are contacted via phone or mail at least two times to minimize missing data.

### Data collection

Questionnaires are provided on paper and are manually entered into the appropriate databases. Measurement data are transferred from the medical device to the institutional server. Additional information regarding disease severity, concomitant diseases and laboratory data are taken from the hospital discharge letter. The vital status is assessed 5 years after study entry or if patients could not be contacted during follow-up.

If patients withdraw consent, all data collected up to the time of revocation will continue to be processed that are necessary for answering the study question. However, patients can actively demand the full deletion of their data.

### Data management and confidentiality

Two staff members independently enter the questionnaire information into the corresponding database. Plausibility ranges for certain variables have been defined during programming of the database, which are automatically checked during data entry. The remaining variables are manually checked for data quality and plausibility. All information from the ACROSSS study is subject to medical confidentiality and data protection regulations. In addition to the local NRW State Data Protection Act, the EU’s General Data Protection Regulation (DSGVO) also applies. Medical and organizational or personal data are kept physically separate in dedicated databases. Only specially authorized staff members have access to both databases to ensure the protection of data. Medical data are stored in pseudonymized form. The manufacturers of the ApneaLink and SOMNOwatch devices are not involved in the study and have no access to the data.

### Confirmation of diagnosis and eligibility criteria

Some patients present with symptoms similar to those of a myocardial infarction, stroke or TIA. Therefore, in addition to admission diagnosis also discharge diagnoses are reexamined in all included patients. If the discharge diagnosis does not match the ICD codes I21, I63, G45.8 or G45.9, patients are excluded. Furthermore, it is checked again in the discharge letter whether it was the first myocardial infarction or stroke and if other exclusion criteria existed.

### Monitoring

The monitoring strategy is adapted to the low risk nature of an observational study. After informed consent is given, completeness of documents is checked by a different staff member. Completeness of questionnaires is checked after each study segment.

## Discussion

The ACROSSS-Study was designed to further investigate the association between sleep characteristics and cardiovascular events. In meta-analyses and systematic reviews, disturbed sleep showed associations with cardiovascular disease, cardiac death and all-cause mortality [40, 41]. However, the ACROSSS study is the first case-crossover study that analyzes if sleep could be an acute trigger for cardiovascular events. If this would be the case, more research would be needed on the physiologic mechanism to find strategies for prevention. Besides the potentially triggering effects of sleep on cardiovascular events, the study focuses also on the health development of the patients after the event. Sleep might modify the process of recovery after the cardiovascular event and could be a potential target to improve this process [42].

A mayor strength of the study is the combination of clinical data from the hospital, objective sleep data, and self-reported information on risk factors and behavior of the patients. For the characterization of sleep, it is important to assess information also for the time before the event, to assess sleep with the same validated questionnaire at the different time points and to combine subjective and objective measurements. The studies published so far included only some of these aspects but not all together [16]. A limitation of the study is that information on sleep habits in several time windows before the myocardial infarction or stroke/TIA is self-reported and misclassification of exposure is likely to occur.

The ACROSSS study will provide comprehensive insights into the sleep characteristics of patients with acute cardiovascular events. Moreover, it offers targets for further causal research on the one hand and targets for a better recovery of patients on the other hand.

## Data availability

All data produced in the present study are available upon reasonable request to the Institute of Medical Informatics, Biometry and Epidemiology, University Hospital Essen, Essen, Germany and requires a contract.

## Ethics approval

This study was performed in line with the principles of the Declaration of Helsinki. Informed consent was obtained from all individual participants included in the study. The conduct of the ACROSSS study has been approved by the central ethics committee of the University of Duisburg-Essen (17-7889-BO) on April 16, 2018. During the course of the study, additional centers were acquired for which ethics votes were obtained from the relevant ethics committees (Medical Council of North-Rhine, Medical Council of Westphalia-Lippe).

## Patient consent

All included patients provided written consent after being thoroughly informed about the study details.

## Trial registration

The ACROSSS study is listed in the German Clinical Trials Register under the ID DRKS00013559 since June 21, 2018.

## Acknowledgement

The authors appreciate the voluntary contribution of all study participants especially with regard to the serious health condition. They also thank the study personnel for their exceptional efforts and the staff of the clinical wards for their support during participant recruitment and taking care of the participants.

## Roles and responsibilities

A. S. and AT. K. compiled the study conception and design. The study is coordinated by AT. K. and T. K.. Patient recruitment is performed by M. C., B. H., B. F., M. K., C. K. and O. B.. The data management is operated by the department for data management at the Institute of Medical Informatics, Biometry and Epidemiology, Essen, Germany. The first draft of the manuscript was written by AT. K. and all authors commented on previous versions of the manuscript. All authors read and approved the final manuscript.

## References

1. World Health Organization. Causes of death. https://www.who.int/news-room/fact-sheets/detail/the-top-10-causes-of-death. Accessed 3 Feb 2021

2. GBD 2019 Diseases and Injuries Collaborators. Global burden of 369 diseases and injuries in 204 countries and territories, 1990–2019: a systematic analysis for the Global Burden of Disease Study 2019. Lancet 396, 1204–1022 (2020).

3. Mahmood, S. S., Levy, D., Vasan, R. S. & Wang, T. J. The Framingham Heart Study and the epidemiology of cardiovascular disease: a historical perspective. Lancet 383, 999–1008 (2014).

4. Kotseva, K. et al. On Behalf Of The Euroaspire Investigators. Lifestyle and risk factor management in people at high risk of cardiovascular disease. A report from the European Society of Cardiology European Action on Secondary and Primary Prevention by Intervention to Reduce Events (EUROASPIRE) IV cross-sectional survey in 14 European regions. Eur. J. Prev. Cardiol. 23, 2007–2018 (2016).

5. Gimbrone, M. A. Jr & García-Cardeña, G. Endothelial cell dysfunction and the pathobiology of atherosclerosis. Circ. Res. 118, 620–636 (2016).

6. Colombo, A., Proietti, R., Culić, V., Lipovetzky, N., Viecca, M. & Danna, P. Triggers of acute myocardial infarction: a neglected piece of the puzzle. J. Cardiovasc. Med. (Hagerstown) 15, 1–7 (2014).

7. Reavey, M., Saner, H., Paccaud, F. & Marques-Vidal, P. Exploring the periodicity of cardiovascular events in Switzerland: variation in deaths and hospitalizations across seasons, day of the week and hour of the day. Int. J. Cardiol. 168, 2195–2200 (2013).

8. Spengos, K. et al. Two-peak temporal distribution of stroke onset in Greek patients. a hospital-based study. Cerebrovasc. Dis. 15, 70–77 (2003).

9. Willich, S. N., Maclure, M., Mittleman, M., Arntz, H. R. & Muller, J. E. Sudden cardiac death. Support for a role of triggering in causation. Circulation 87, 1442–1450 (1993).

10. Johansson, I., Karlson, B. W., Grankvist, G. & Brink, E. Disturbed sleep, fatigue, anxiety and depression in myocardial infarction patients. Eur. J. Cardiovasc. Nurs. 9, 175–180 (2010).

11. Baylor, C., Yorkston, K. M., Jensen, M. P., Truitt, A. R. & Molton, I. R. Scoping review of common secondary conditions after stroke and their associations with age and time post stroke. Top. Stroke Rehabil. 21, 371–382 (2014).

12. Alcantara, C., Peacock, J., Davidson, K. W., Hiti, D. & Edmondson, D. The association of short sleep after acute coronary syndrome with recurrent cardiac events and mortality. Int. J. Cardiol. 171, e11–2; 10.1016/j.ijcard.2013.11.135 (2014).

13. Coryell, V. T., Ziegelstein, R. C., Hirt, K., Quain, A., Marine, J. E. & Smith, M. T. Clinical correlates of insomnia in patients with acute coronary syndrome. Int. Heart J. 54, 258–265 (2013).

14. BaHammam, A. Sleep quality of patients with acute myocardial infarction outside the CCU environment: a preliminary study. Med. Sci. Monit. 12, CR168–172 (2006).

15. Schiza, S. E. et al. Sleep patterns in patients with acute coronary syndromes. Sleep Med. 11, 149–153 (2010).

16. Madsen, M. T., Huang, C., Zangger, G., Zwisler, A. D. O. & Gögenur, I. Sleep disturbances in patients with coronary heart disease: A systematic review. J. Clin. Sleep Med. 15, 489–504 (2019).

17. Mittleman, M. A., Maclure, M., Tofler, G. H., Sherwood, J. B., Goldberg, R. J. & Muller, J. E. Triggering of acute myocardial infarction by heavy physical exertion. Protection against triggering by regular exertion. Determinants of Myocardial Infarction Onset Study Investigators. N. Engl. J. Med. 329, 1677–1683 (1993).

18. Maclure, M. & Mittleman, M. A. Should we use a case-crossover design? Annu. Rev. Public Health 21, 193–221 (2000).

19. Stang, A. et al. Daily siesta, cardiovascular risk factors, and measures of subclinical atherosclerosis: results of the Heinz Nixdorf Recall Study. Sleep 30, 1111–1119 (2007).

20. German Stroke Aid Foundation. Symptoms of stroke. https://www.schlaganfall-hilfe.de/de/verstehen-vermeiden/schlaganfall-erkennen/symptome. Accessed Oct 2017

21. German Heart Foundation. Symptoms of myocardial infarction. https://www.herzstiftung.de/infos-zu-herzerkrankungen/herzinfarkt/anzeichen. Accessed Oct 2017

22. Lipovetzky, N., Hod, H., Roth, A., Kishon, Y., Sclarovsky, S. & Green, M. S. Heavy meals as a trigger for a first event of the acute coronary syndrome: a case-crossover study. Isr. Med. Assoc. J. 6, 728–731 (2004).

23. Baylin, A., Hernandez-Diaz, S., Kabagambe, E. K., Siles, X. & Campos, H. Transient exposure to coffee as a trigger of a first nonfatal myocardial infarction. Epidemiology 17, 506–511 (2006).

24. Gerlich, M. G. et al. Patterns of alcohol consumption and acute myocardial infarction: a case-crossover analysis. Eur. Addict. Res. 15, 143–149 (2009).

25. Wen, Y. C., Hsiao, F. Y., Chan, K. A., Lin, Z. F., Shen, L. J. & Fang, C. C. Acute respiratory infection and use of nonsteroidal anti-inflammatory drugs on risk of acute myocardial infarction: a nationwide case-crossover study. J. Infect. Dis. 215, 503–509 (2017).

26. Koton, S., Tanne, D., Bornstein, N. M. & Green, M. S. Triggering risk factors for ischemic stroke: a case-crossover study. Neurology 63, 2006–2010 (2004).

27. Smyth, A. et al.; INTERHEART Investigators. Physical activity and anger or emotional upset as triggers of acute myocardial infarction: The INTERHEART study. Circulation 134, 1059–1067 (2016).

28. von Klot, S. et al. Intensity of physical exertion and triggering of myocardial infarction: a case-crossover study. Eur. Heart J. 29, 1881–1888 (2008).

29. Buysse, D. J., Reynolds, C. F. 3rd, Monk, T. H., Berman, S. R. & Kupfer, D. J. The Pittsburgh Sleep Quality Index: a new instrument for psychiatric practice and research. Psychiatry Res. 28, 193–213 (1989).

30. Krotsetis, S., Richards, K. C., Behncke, A. & Köpke, S. The reliability of the German version of the Richards Campbell Sleep Questionnaire. Nurs. Crit. Care 22, 247–252 (2017).

31. Netzer, N. C., Stoohs, R. A., Netzer, C. M., Clark, K. & Strohl, K. P. Using the Berlin Questionnaire to identify patients at risk for the sleep apnea syndrome. Ann Intern Med 131, 485–491 (1999).

32. Johns, M. W. A new method for measuring daytime sleepiness: the Epworth sleepiness scale. Sleep 14, 540–545 (1991).

33. Schmermund, A. et al. Assessment of clinically silent atherosclerotic disease and established and novel risk factors for predicting myocardial infarction and cardiac death in healthy middle-aged subjects: rationale and design of the Heinz Nixdorf RECALL Study. Risk factors, evaluation of coronary calcium and lifestyle. Am. Heart J. 144, 212–218 (2002).

34. Peters, A. et al., German National Cohort (GNC) Consortium. Framework and baseline examination of the German National Cohort (NAKO). Eur. J. Epidemiol. 37, 1107–1124 (2022).

35. Greenland, S., Pearl, J. & Robins, J. M. Causal diagrams for epidemiologic research. Epidemiology 10, 37–48 (1999).

36. Bassetti, C. L. A. et al. EAN/ERS/ESO/ESRS statement on the impact of sleep disorders on risk and outcome of stroke. Eur. Respir. J. 55, 1901104; 10.1183/13993003.01104-2019 (2020).

37. Erman, M. K., Stewart, D., Einhorn, D., Gordon, N. & Casal, E. Validation of the ApneaLink for the screening of sleep apnea: a novel and simple single-channel recording device. J. Clin. Sleep Med. 3, 387–392 (2007).

38. Weinreich, G., et al; Heinz Nixdorf Recall (HNR) study group. Association of obstructive sleep apnea with subclinical coronary atherosclerosis. Atherosclerosis 231, 191–197 (2013).

39. Zinkhan, M. et al. Agreement of different methods for assessing sleep characteristics: a comparison of two actigraphs, wrist and hip placement, and self-report with polysomnography. Sleep Med. 15, 1107–1114 (2014).

40. Sofi, F., Cesari, F., Casini, A., Macchi, C., Abbate, R. & Gensini, G. F. Insomnia and risk of cardiovascular disease. A meta-analysis. Eur. J. Prev. Cardiol. 21, 57–64 (2014).

41. Cappuccio, F. P., Cooper, D., D’Elia, L., Strazzullo, P. & Miller, M. A. Sleep duration predicts cardiovascular outcomes. A systematic review and meta-analysis of prospective studies. Eur. Heart J. 32, 1484–1492 (2011).

42. Rouleau, C. R., Toivonen, K., Aggarwal, S., Arena, R. & Campbell, T. S. The association between insomnia symptoms and cardiovascular risk factors in patients who complete outpatient cardiac rehabilitation. Sleep Med. 32, 201–207 (2017).

